# Plasma Metabolomics During Chemotherapy Desensitization: A Pilot Study

**DOI:** 10.64898/2026.10.30.26366681

**Authors:** Amir Asiaee, Stacy D. Sherrod, Simona G. Codreanu, Katrina L. Leaptrot, Alexandra C. Schrimpe-Rutledge, John A. McLean, Micah A. McGinnis, Halle C. Petrie, Lauren S. Prescott, Alaina J. Brown, Ronald Alvarez, Elizabeth J. Phillips, Cosby A. Stone

## Abstract

**Background:** Rapid drug desensitization permits continued chemotherapy, but no validated blood measurement predicts breakthrough reactions during desensitization.

**Objective:** To compare paired plasma metabolomic and lipidomic changes during chemotherapy desensitization with those in tolerant controls, and to describe breakthrough and omalizumab encounters.

**Methods:** In this pilot study, 10 patients undergoing carboplatin or paclitaxel desensitization and four tolerant controls receiving standard infusions contributed 76 draws across 39 encounters (37 with paired pre- and post-infusion draws). Untargeted liquid chromatography-mass spectrometry in four modes yielded 8,646 quality-filtered features (instrument signals). Exact permutation with false discovery rate control compared patient-averaged paired changes from 27 allergic encounters (10 patients) off omalizumab with four control encounters (four patients). Breakthrough and omalizumab comparisons were descriptive.

**Results:** Allergic patients’ profiles changed from pre- to post-infusion in three of four modes (Holm-adjusted *P* = .04). Pre-infusion profiles differed between allergic patients and controls in one mode (Holm-adjusted *P* = .02). Montelukast, a premedication drug, had the smallest *P* value among annotated pre-infusion differences and was detected in all 33 allergic and no control pre-infusion draws. No feature distinguished allergic from control paired changes at a 5% false discovery rate. Histamine did not consistently increase in seven paired breakthrough encounters (five patients). Hemolysis was more frequent in allergic post-infusion than pre-infusion draws (16 of 35 vs eight of 33).

**Conclusion:** Profiling during desensitization detected premedication exposure and broad pre-to-post changes largely shared with controls, but no allergy-specific signature. Prospective studies need sampling timed to reactions and medications, comparators matched for premedication and infusion duration, and standardized specimen handling.

**Highlights Box:** *What is already known about this topic?:* Desensitization lets most patients with carboplatin or paclitaxel hypersensitivity continue treatment. Breakthrough reactions occur, and no validated blood measurement predicts them.

*What does this article add to our knowledge?:* In 14 patients, plasma profiling detected premedication signals before infusion and broad pre-to-post changes; no feature distinguished allergic from control changes at a 5% false discovery rate. Histamine did not consistently rise with breakthrough reactions.

*How does this study impact current management guidelines?:* Biomarker studies during chemotherapy desensitization should match comparators for premedication, time blood draws to symptom onset, and standardize specimen processing; these exploratory findings do not change current desensitization practice.

## Introduction

Carboplatin and paclitaxel remain the backbone of treatment for ovarian and endometrial cancer. Immediate hypersensitivity reactions to both agents are common;^1,2^ roughly one fifth to one half of patients re-treated with carboplatin react, depending on the platinum-free interval.^3,4^ Rapid drug desensitization allows most affected patients to continue the planned regimen,^5,6,7^ with breakthrough symptoms in roughly one in 10 courses.^8^ Case reports and small case series describe adding omalizumab, an anti-IgE antibody, for patients with breakthrough reactions during desensitization or severe initial reactions.^9,10,11,12^ After a breakthrough reaction, the allergist decides whether to lengthen the protocol, intensify premedication, or add omalizumab on clinical grounds alone. The same immediate reaction can arise through different mechanisms, so the clinical picture alone does not show which one is at work. IgE activates mast cells and basophils in immediate allergic reactions,^13^ and IgE-dependent basophil activation has been shown in severe carboplatin reactions,^14^ whereas other drugs activate mast cells without IgE through Mas-related G protein-coupled receptor X2 and produce similar reactions.^15^ No validated blood measurement predicts breakthrough during desensitization.

Untargeted metabolomics measures thousands of small molecules and lipids in a single plasma sample. A 2024 systematic review found a single human metabolomic study of anaphylaxis, in which changes in phospholipids, glucose, and cortisol differed by trigger and severity;^16,17^ arachidonic acid, tryptophan, and lysophosphatidylcholines were altered across the animal and human studies reviewed.^16^ Prior metabolomic studies of drug hyper-sensitivity profiled plasma in liposomal doxorubicin infusion reactions, including pre-infusion samples,^18,19^ and around aspirin desensitization;^20^ we found none that sampled plasma serially during platinum or taxane desensitization. Paired sampling before and after an infusion removes stable between-person differences, but the infusion itself, premedication given to allergic patients, and specimen handling^21,22^ can still alter measured profiles. A comparator group receiving the same chemotherapy by standard infusion separates changes shared by both groups from those specific to desensitized patients.

We performed paired plasma metabolomic and lipidomic profiling in 10 patients undergoing chemotherapy desensitization after an immediate hypersensitivity reaction to carboplatin or paclitaxel and in four *tolerant controls*, patients receiving the same chemotherapy by standard infusion. The primary question was whether pre-to-post changes differed between allergic patients and controls. We also compared paired changes between breakthrough and non-breakthrough desensitization encounters and between encounters on and off omalizumab in its two recipients, and we examined premedication signals, reaction sampling, and specimen quality.

## Methods

### Study design and participants

This prospective observational study enrolled patients in the chemotherapy desensitization program at Vanderbilt University Medical Center under a protocol approved by its Institutional Review Board (#221684). All participants gave written informed consent, and the study was conducted in accordance with the Declaration of Helsinki. The allergic group comprised 10 patients with a prior immediate hypersensitivity reaction to carboplatin (six), paclitaxel (three), or both (one) who subsequently received the culprit agent by rapid drug desensitization. Allergy classification and the decision to desensitize were based on the clinical reaction and specialist assessment; chemotherapy skin testing was not routinely performed. The control group comprised four patients who tolerated standard carboplatin-paclitaxel infusions with standard chemotherapy premedication and were sampled at varying treatment cycles; unlike standard infusions, desensitization infusions used extended multi-step administration and included meals. Age, sex, race, ethnicity, cancer type and stage, and *BRCA* status were obtained from electronic health record review and the enrollment survey.

### Desensitization, premedication, and sampling

Desensitization followed the program’s pharmacy-prepared three-bag, 13-step protocol,^7^ adapted from Castells and colleagues’ 12-step protocol.^5,8^ Premedication comprised cetirizine or fexofenadine, famotidine, and montelukast from two days before through two days after desensitization, with standard chemotherapy premedication when indicated.^7^ An encounter is one sampled infusion date, and a breakthrough reaction is a recorded hypersensitivity reaction during desensitization. Pre draws were obtained before the start of paclitaxel and carboplatin administration. Post draws were collected after infusion completion or, in breakthrough encounters, as soon as possible after the reaction was treated; 37 encounters had assayed pre and post draws, and two breakthrough encounters had post draws only.

Omalizumab exposure at each draw was determined from study records and administration dates; both omalizumab recipients started treatment after a breakthrough reaction. Plasma was obtained during automated cell separation (RoboSep) or density-gradient separation in Ficoll barrier tubes, stored frozen, and analyzed in a single batch. Hemolysis was assessed visually, and post draws processed on the calendar day after collection were classified as next-day processed. Specimen processing is described in the Online Repository.

### Metabolomic and lipidomic measurements

The Vanderbilt Center for Innovative Technology analyzed 60-*µ*L plasma aliquots by untargeted liquid chromatography-tandem mass spectrometry (LC-MS/MS) in four modes: hydrophilic interaction liquid chromatography (HILIC) with positive and negative ionization and reversed-phase liquid chromatography (RPLC) with negative ionization for metabolites on a Thermo Scientific Q Exactive HF Orbitrap, and RPLC with negative ionization for lipids on an Agilent 6560 quadrupole time-of-flight (Q-TOF) mass spectrometer. A pooled quality control sample, prepared by mixing equal volumes of all study plasma samples and injected repeatedly within each of the four sample sets, and isotope-labeled internal standards were used to monitor reproducibility. Each feature represents a unique retention time and mass-to-charge ratio; a single compound can yield several features. An annotation is a proposed chemical identity graded on a five-level scheme adapted from the Metabolomics Standards Initiative, from confirmed identification (Level 1) through putative (Level 2), tentative (Level 3), and molecular-formula (Level 4) annotations to unknown features (Level 5).^23,24^ We retained features with no more than 20% zero or missing study-sample values and a coefficient of variation of at most 30% across the seven injections of the pooled quality control sample, replaced remaining zeros with half the feature’s minimum positive value, and analyzed log_2_ relative peak areas. Of 10,235 features, 8,646 were retained, including 45 with Level 1 and 477 with Level 2 annotations; sample preparation, instrumentation, and annotation are detailed in the Online Repository.

### Statistical analysis

We computed each feature’s paired change as log_2_(post/pre) within an encounter. The primary comparison included 27 encounters from 10 allergic patients and four encounters from four controls, all off omalizumab at both draws (see Table E1 in the Online Repository). We averaged paired changes across eligible encounters within each patient and compared group means, with equal patient weights, by a two-sided exact test that enumerated all 1,001 assignments of four control labels among the 14 patient means; its smallest attainable *P* value was therefore 1/1,001.^25^ As a pilot study, enrollment was not based on a formal sample-size calculation. We applied Benjamini-Hochberg adjustment within mode at a 5% false discovery rate (FDR),^26^ and Welch *t* tests supplied pointwise 95% confidence intervals (CIs).^27^ Sensitivity analyses repeated the comparison with seven alternative methods, including a random-intercept mixed model of encounter-level changes,^28^ empirical-Bayes moderated tests, and Storey *q* values (Online Repository).

We tested feature-level pre-to-post changes within allergic patients by exact sign-flip permutation of patient means and pre-infusion differences between allergic patients and controls by exact patient-label permutation, each with Benjamini-Hochberg adjustment within mode. To assess whether an assay’s overall profile differed between groups or changed within groups, we used mode-wide global tests with the same exact patient-label and sign-flip permutations.^29^ Holm adjustment covered 20 global tests, five questions in each of four modes: allergic versus control change, pre-infusion difference, carboplatin-restricted change, and pre-to-post change within allergic patients and within controls.^30^ Breakthrough versus non-breakthrough contrasts were computed within the two patients observed in both states off omalizumab, and omalizumab contrasts within each recipient; both were descriptive. Medication, histamine, bile-acid, and principal-component analyses were post hoc. Analyses used R 4.6.1 (R Foundation for Statistical Computing, Vienna, Austria) with nlme 3.1-169, limma 3.68.5, and qvalue 2.44.0.

## Results

### Patients, encounters, and specimens

Between October 2024 and October 2025, 14 patients contributed 76 draws from 39 infusion encounters: 37 complete pairs (33 allergic, four control) and two post-only reaction draws (Table I; Figure 1; Figure E1 in the Online Repository). All 14 patients were women and non-Hispanic. Allergic patients were in their 40s to 60s and had ovarian (six) or endometrial (four) cancer; controls were in their 50s to 70s and had squamous cell carcinoma of the buccal mucosa, lung, or skin (three) or endometrial cancer (one). Eight allergic patients were White and two were Black; three controls were White and one was Black. Allergic patients had one to seven encounters each, and controls contributed one pair each. The primary comparison used 27 allergic pairs off omalizumab and four control pairs. Nine breakthrough encounters occurred in five patients, all off omalizumab. Patients A07 (carboplatin) and A09 (paclitaxel) began omalizumab after breakthrough and contributed six on-treatment pairs with no recorded reaction.

**Table I:** Patient characteristics and sampled encounters.

| Patient | Group, culprit drug | Age group, y | Cancer, stage | BRCA | Encounters, paired (post only) | Breakthrough encounters | Omalizumab pairs | Hemolyzed/total draws |
| --- | --- | --- | --- | --- | --- | --- | --- | --- |
| A01 | Allergic, paclitaxel | 50–54 | Endometrial, II | No | 1 (0) | 1 | 0 | 0/2 |
| A02 | Allergic, carboplatin | 40–44 | Ovarian, IIIC | *BRCA1* | 5 (0) | 0 | 0 | 2/10 |
| A03 | Allergic, carboplatin | 60–64 | Endometrial, IB | No | 3 (0) | 0 | 0 | 0/6 |
| A04 | Allergic, carboplatin | 60–64 | Ovarian, IV | No | 4 (0) | 1 | 0 | 2/8 |
| A05 | Allergic, carboplatin | 40–44 | Ovarian, IIIC | No | 3 (1) | 4 | 0 | 5/7 |
| A06 | Allergic, carboplatin | 55–59 | Ovarian, IV | No | 1 (0) | 0 | 0 | 0/2 |
| A07 | Allergic, carboplatin | 60–64 | Ovarian, IV | No | 7 (0) | 1 | 3 | 5/14 |
| A08 | Allergic, both | 60–64 | Ovarian, IIIA2 | *BRCA2* | 3 (0) | 0 | 0 | 6/6 |
| A09 | Allergic, paclitaxel | 65–69 | Endometrial, IV | No | 4 (1) | 2 | 3 | 3/9 |
| A10 | Allergic, paclitaxel | 50–54 | Endometrial, IA | No | 2 (0) | 0 | 0 | 1/4 |
| T01 | Tolerant control | 75–79 | SCC of buccal mucosa, I | *BRCA2* | 1 (0) | 0 | 0 | 0/2 |
| T02 | Tolerant control | 60–64 | SCC of lung, IV | *BRCA1* | 1 (0) | 0 | 0 | 2/2 |
| T03 | Tolerant control | 50–54 | SCC of skin, III | Not tested | 1 (0) | 0 | 0 | 0/2 |
| T04 | Tolerant control | 55–59 | Endometrial, IIIB | No | 1 (0) | 0 | 0 | 0/2 |

Clinical fields come from chart review and the enrollment survey. Patient codes (A01 to A10, allergic; T01 to T04, tolerant controls) were assigned for this report and are not linked to medical record or study enrollment numbers. Ages are given in 5-year groups. All 14 patients were women and non-Hispanic. Controls received standard carboplatin-paclitaxel infusions. Culprit drug is the agent implicated in the prior reaction; both denotes carboplatin and paclitaxel. Encounters are sampled infusion dates; counts give complete pre/post pairs, with post-only reaction draws in parentheses. Hemolyzed/total draws gives draws with visually noted hemolysis over all draws for that patient. *BRCA*, breast cancer susceptibility gene (*BRCA1* or *BRCA2*) pathogenic variant; SCC, squamous cell carcinoma.

**Figure 1:**
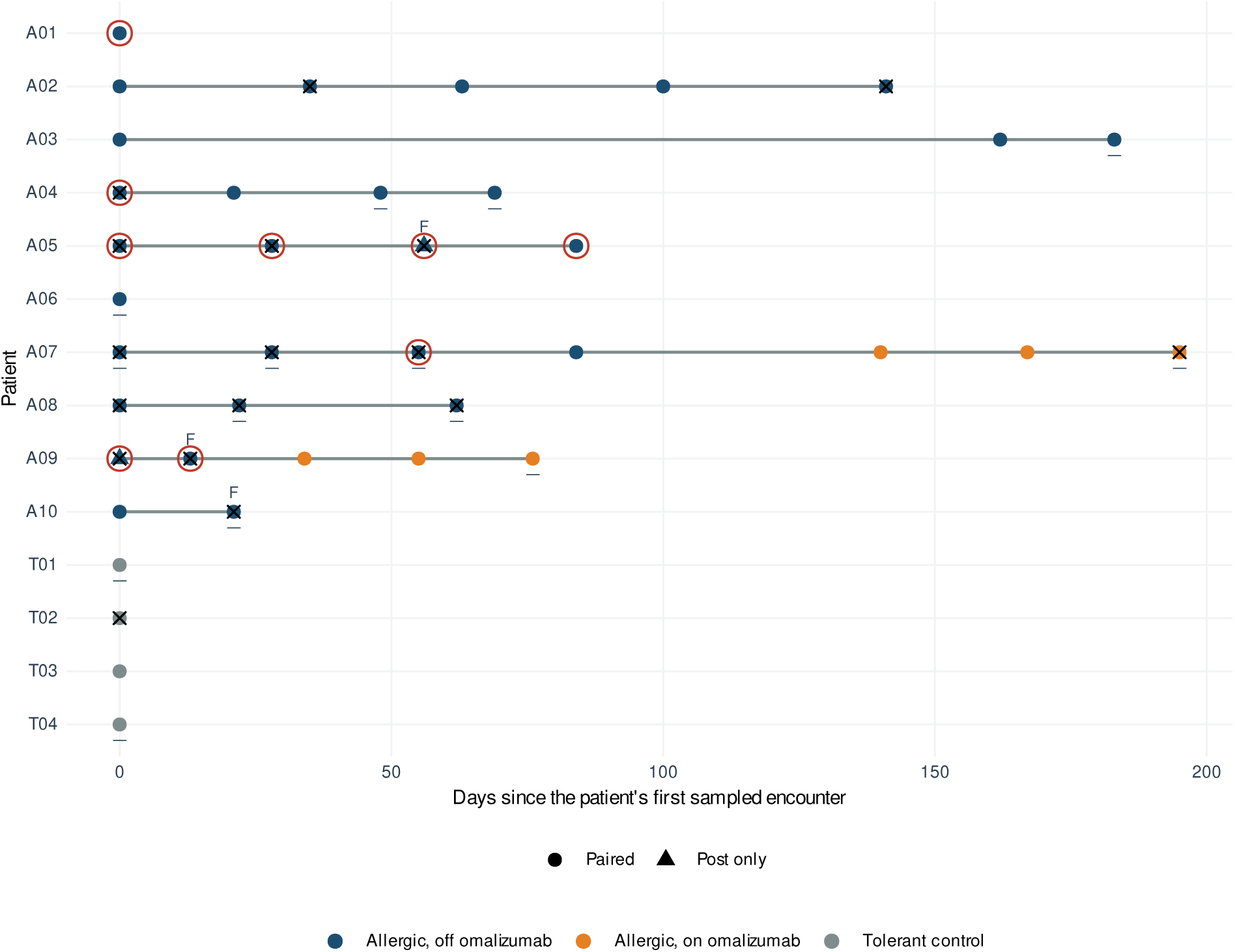
Sampled encounters for all 14 patients. Points represent infusion encounters: circles, complete pre/post pairs; triangles, post draws only. Colors indicate group and omalizumab status. The horizontal axis gives days since each patient’s first sampled encounter. Red rings mark breakthrough reactions; crosses, recorded hemolysis; bars below points, next-day processing of post draws; F, Ficoll/barrier-tube processing. Lines connect each patient’s encounters.

Hemolysis was noted in 26 draws: 16 of 35 allergic post draws, eight of 33 allergic pre draws, and two of eight control draws; seven of the nine breakthrough post draws were hemolyzed (Figure E2 in the Online Repository). Twelve allergic and two control post draws, and no pre draws, were processed the next calendar day.

### Pre-infusion differences and premedication

The pre-infusion comparison averaged each patient’s pre draws from primary-comparison encounters (27 draws from 10 allergic patients; four from four controls). Profiles differed globally in HILIC-positive mode (exact *P* = .001, the minimum attainable; Holm-adjusted *P* = .02) but not in HILIC-negative, RPLC-negative, or lipid-negative mode after Holm adjustment (unadjusted *P* = .02, .17, and .14, respectively; Table E6). No individual pre-infusion feature met the 5% FDR threshold (Figure E3); among annotated features, montelukast had the smallest Welch *P* value. In a post hoc analysis of two premedication annotations, montelukast (Level 2) was detected in all 33 allergic pre-infusion draws, 34 of 35 allergic post-infusion draws, and none of the eight control draws. Its signal fell from pre- to post-infusion in 32 of 33 paired allergic encounters and rose in one (median, *−*1.9 log_2_; interquartile range, *−*2.7 to *−*1.2; Figure 2; Table E2). Cetirizine (Level 2) was detected in 31 of 33 allergic pre draws, 31 of 35 allergic post draws, and two of four control draws at each time point.

**Figure 2:**
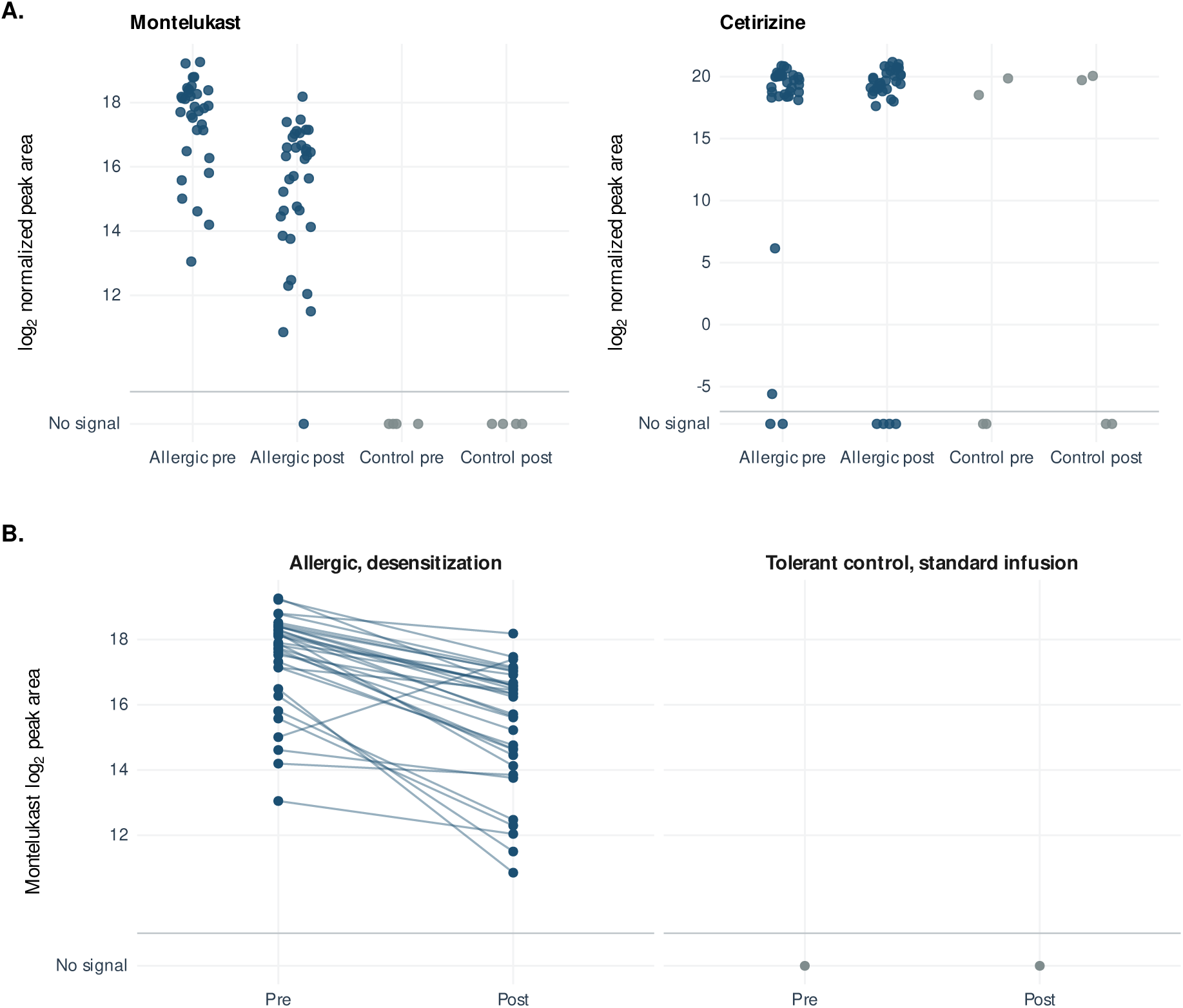
Premedication annotations by group and draw. A. Montelukast and cetirizine in all 76 draws; zeros appear in the separate “No signal” row. B. Montelukast in 37 complete encounters; lines connect pre and post draws with signal at both. Values are relative log_2_ peak areas in HILIC-positive mode. HILIC, hydrophilic interaction liquid chromatography.

### Pre-to-post change within allergic patients

In allergic patients off omalizumab (27 pairs from 10 patients), the global sign-flip permutation test attained its minimum *P* value in HILIC-positive, HILIC-negative, and RPLC-negative modes (*P* = .002; Holm-adjusted *P* = .04) and gave *P* = .004 in lipid-negative mode (Holm-adjusted *P* = .06; Table E6). Of the 1,070 features that changed at a 5% within-allergic FDR (129, 609, 78, and 254 in those four modes, respectively), 58% to 94% changed in the same direction as the mean control change. Across features, the allergic and control mean paired changes were positively correlated across modes (*r* = 0.54 to 0.85). Table E3 lists the 36 Level 1 and Level 2 annotations among the within-allergic changes.

### Allergic versus tolerant patients

No feature distinguished allergic from control paired changes at a 5% FDR in any mode, by the exact test or by any of the seven alternative methods (Table E4 and Figure E4); the smallest adjusted *P* value was .07. The number of features with unadjusted Welch *P < .*05 exceeded the number expected under the null hypothesis in HILIC-negative (167 vs 151) and RPLC-negative (141 vs 93) modes, but not in HILIC-positive (125 vs 125) or lipid-negative (57 vs 64) modes (Figure 3, A). Mode-wide global *P* values were .15 in HILIC-positive, .054 in HILIC-negative, .09 in RPLC-negative, and .57 in lipid-negative mode; none was significant after Holm adjustment (Table E6). The annotated features with the smallest Welch *P* values were montelukast (difference in paired change, *−*1.8 log_2_; 95% CI, *−*2.4 to *−*1.3) and traumatic acid, a plant-derived dicarboxylic acid (0.9; 95% CI, 0.6 to 1.2), followed by lysophosphatidylcholine and lysophosphatidylethanolamine species, citrate, and saccharopine (Figure 3, B; 42 features outside the *±*5 log_2_ display range are shown in Figure E5). A feature annotated as sedoheptulose 7-phosphate (Level 3), an intracellular pentose phosphate pathway intermediate, ranked seventh by Welch *P* in HILIC-negative mode (2.4; 95% CI, 1.2 to 3.5).

**Figure 3:**
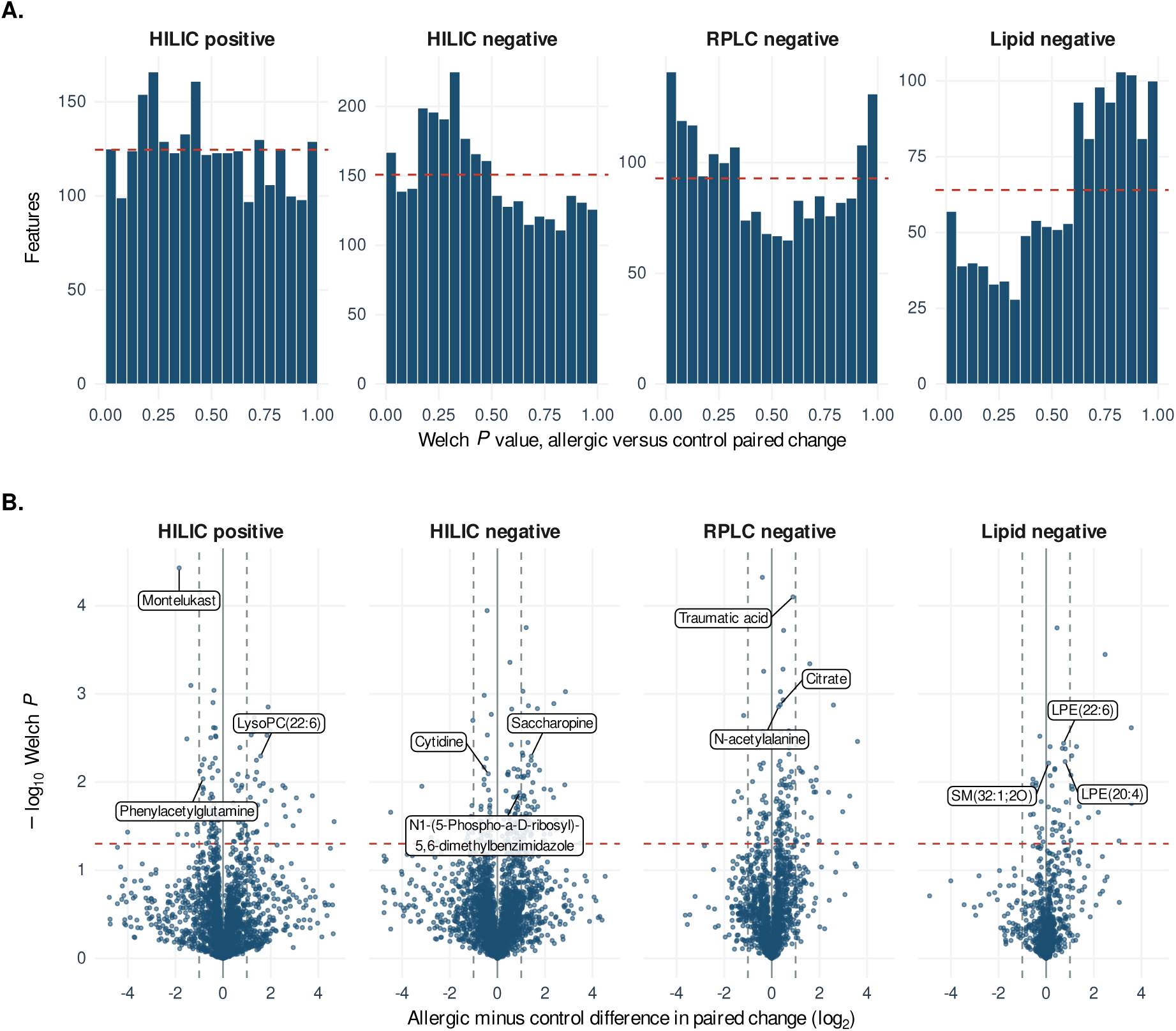
Allergic versus tolerant difference in paired change. A. Welch *P* values by mode; dashed lines show uniform expectations. B. Paired-change differences against Welch *P* values; dashed lines mark twofold differences and nominal *P* = .05. Labels mark the three smallest-*P* Level 1 or 2 features per mode. HILIC, hydrophilic interaction liquid chromatography; RPLC, reversed-phase liquid chromatography.

At a 10% FDR, few features were selected, and the selections differed by method (Table E4). Analyses restricted to pairs without hemolysis, with the same hemolysis status at both draws, with RoboSep processing, or with same-day processing, and analyses with stricter missing-value filters also yielded no discoveries at a 5% FDR (Table E5 and Online Repository). Among the 21 features whose top-ranked annotation was a bile acid or bile-acid conjugate, 18 had more positive paired changes in allergic patients than in controls. Patient-mean paired changes averaged across this set were +0.59 log_2_ in allergic patients versus *−*0.49 in controls; one feature’s pointwise 95% CI excluded zero (Figure E6). Principal components of the 14 patient-mean change vectors did not separate the groups (Figure E7).

### Breakthrough encounters and histamine

Seven of the nine breakthrough encounters had paired pre- and post-infusion draws. Within-patient comparison was possible in patients A04 and A07, who together contributed two breakthrough and six non-breakthrough pairs off omalizumab. Differences between break-through and non-breakthrough mean paired changes had the same sign in the two patients for 38% to 68% of features across modes (50% expected by chance). Median post-infusion histamine (Level 1) was 12.6 log_2_ relative peak area in control draws (four patients), 12.6 in non-breakthrough allergic draws (26 draws, eight patients), and 12.5 in breakthrough draws (nine draws, five patients). Median histamine paired change was *−*0.01 (range, *−*0.66 to 0.29) across all seven breakthrough pairs from five patients, 0.04 (*−*0.74 to 0.67) across 26 non-breakthrough allergic pairs from eight patients, including six on omalizumab, and 0.05 (*−*0.08 to 0.38) across the four control pairs (Figure 4, A and B).

**Figure 4:**
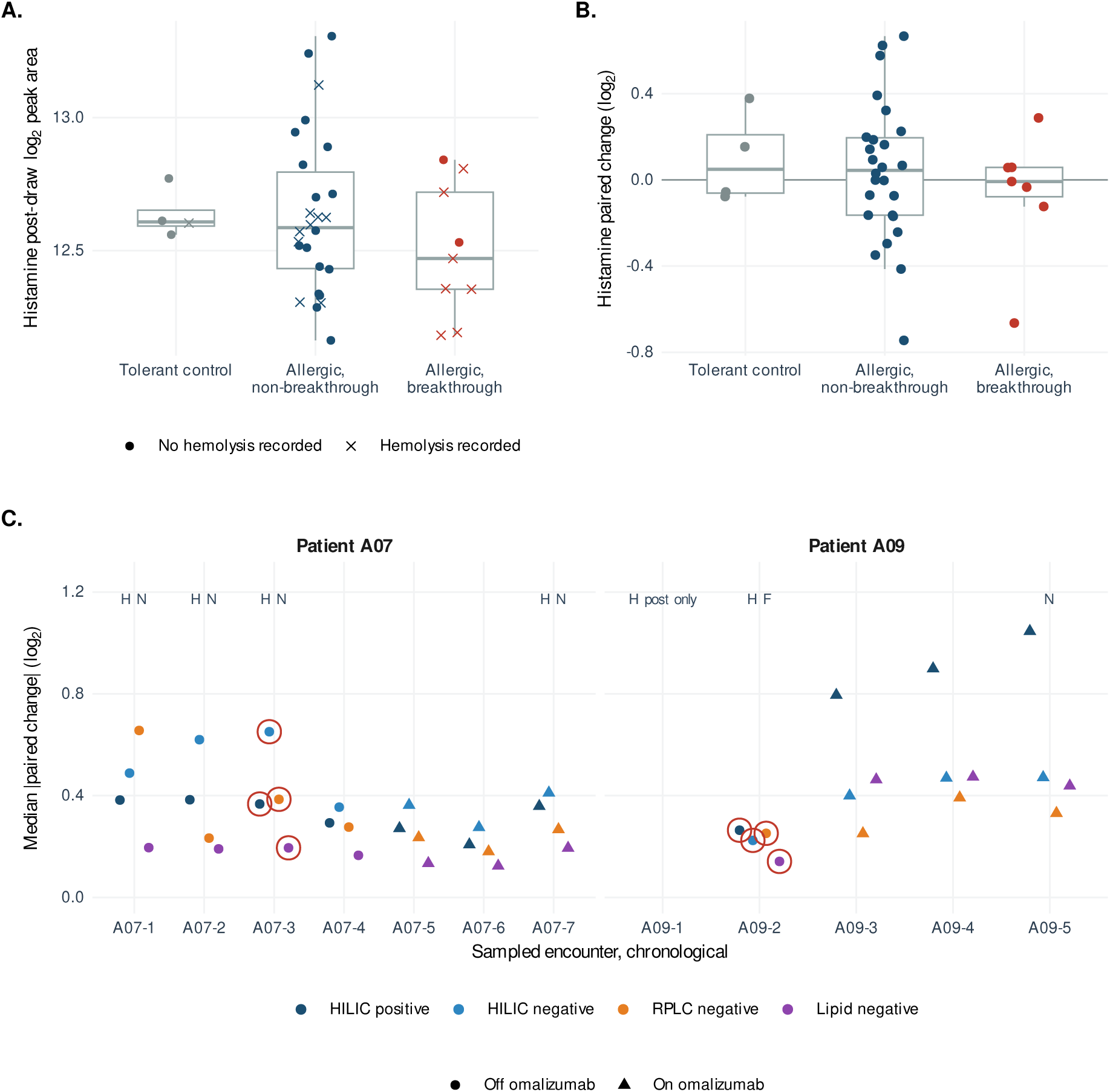
Histamine by encounter and paired changes in omalizumab recipients. A. Post-draw histamine; crosses mark hemolysis. B. Histamine paired change in complete encounters. C. Median absolute paired change by mode for patients A07 and A09. Red rings mark breakthrough; H, hemolysis; N, next-day processing; F, Ficoll/barrier-tube processing. HILIC, hydrophilic interaction liquid chromatography; RPLC, reversed-phase liquid chromatography.

### Omalizumab recipients

Patient A07 contributed four off-treatment and three on-treatment pairs; patient A09 contributed one off-treatment and three on-treatment pairs plus an earlier post-only reaction draw (Figure 4, C). At each encounter, the median absolute paired change across all features summarized the magnitude of profile change in log_2_ units. It was similar on and off treatment in patient A07 (HILIC positive, 0.29 to 0.38 off and 0.21 to 0.36 on) and larger on treatment in patient A09 (0.26 off and 0.79 to 1.05 on). Patient A09’s single off-treatment pair was hemolyzed and Ficoll processed, and two of the patient’s on-treatment post-infusion draws had about five times the median number of zero values (see the Online Repository). For each patient and feature, we subtracted the mean off-omalizumab paired change from the mean on-omalizumab paired change. These differences agreed in sign between the two patients for 40% to 59% of features across modes (50% expected by chance; Figure E8).

## Discussion

We found that plasma profiling during chemotherapy desensitization captured a broad pre-to-post shift in allergic patients, mostly in the same direction as the change in tolerant controls, and a pre-infusion difference from controls in one assay mode. No individual feature distinguished allergic from control paired changes, by the primary exact test or by seven alternative methods; the smallest adjusted *P* value was .07. The excess of small Welch *P* values in HILIC-negative and RPLC-negative modes is compatible with modest group differences that a cohort of 14 patients cannot resolve feature by feature.

The clearest difference between groups reflected a premedication drug: montelukast, given only to allergic patients before desensitization, was detected in all 33 allergic pre-infusion draws and in no control draw, had the strongest pre-infusion difference among annotated features, and may have contributed to the global difference in HILIC-positive mode. Cetirizine was also detected in two of four controls, so the comparator group was not antihistamine-free. Because its signal fell during the visit, paired subtraction does not remove it from pre-to-post comparisons. The other annotated feature with a small Welch *P* value, traumatic acid, is a plant-derived compound that more plausibly reflects diet than host biology. Future studies should therefore record medications and recent meals at each draw, or match comparators on them.

Earlier metabolomic studies of drug hypersensitivity were similarly small. In plasma drawn before PEGylated liposomal doxorubicin infusions in 14 patients, L-histidine was higher and urocanic acid lower in those who had immediate reactions,^18^ and a 12-patient pilot of liposomal doxorubicin reported near-perfect discrimination by several metabolites.^19^ In aspirin desensitization, plasma salicylic acid was lower in patients than in controls before desensitization,^20^ a drug-exposure difference comparable to our montelukast signal. In serum sampled during and after anaphylaxis, metabolic changes differed by trigger and severity.^17^ Patients who react and those who do not can differ in premedication and other exposures, and our data show that medication exposure alone can produce a group difference larger and more consistent than any host signal. Candidate markers reported from small cohorts therefore need replication with comparators matched for medication exposure.

Phospholipids changed during anaphylaxis in the one human study, and lysophosphatidylcholines were among the metabolites altered across anaphylaxis models;^16,17^ lysophosphatidylethanolamines and a lysophosphatidylcholine were among our lipid features with small unadjusted *P* values. The average paired change across 21 bile-acid features was positive in allergic patients and negative in controls. These patterns could reflect hypersensitivity-related biology or the surrounding care: desensitization infusions were longer, included meals, which can contribute to bile-acid changes, and added premedication. The Level 3 sedoheptulose 7-phosphate annotation also had a small unadjusted *P* value; because it is an intracellular metabolite, its release during hemolysis, which was more frequent in allergic post draws, offers an alternative explanation. Hemolysis and delayed processing are known to alter plasma metabolomic profiles^21,22^ and may contribute to these differences.

Histamine (Level 1) did not consistently increase in paired breakthrough samples. Because circulating histamine changes rapidly, sampling after the peak can miss a transient rise,^31,32^ and draws timed to symptom onset are needed to distinguish a missed peak from a lack of histamine release. Neither omalizumab recipient had a recorded breakthrough reaction in six on-treatment encounters. Because only two patients received omalizumab, both starting after a breakthrough reaction, and specimen quality differed between their on- and off-treatment draws, these observations are descriptive.

These observations inform sampling in prospective studies of desensitization. First, pre-medication is a measurable exposure that changes during the visit, so candidate markers need either comparators matched for premedication and infusion duration or direct measurement and modeling of medication exposure. Second, evaluating transient markers of breakthrough reactions requires, at minimum, a timed draw soon after symptom onset, without delaying rescue treatment, and recorded rescue medication times. Third, seven of nine breakthrough post draws were hemolyzed, compared with nine of 26 non-breakthrough allergic post draws, which may reflect chemotherapy exposure, processing time, or both. Standardized processing,^33^ quantitative hemolysis assessment, and documented meals and medications are needed before differences can be attributed to the reaction.

To our knowledge, this is the first study to profile plasma serially by untargeted metabolomics during platinum or taxane desensitization. The design paired pre and post draws within each encounter, gave controls the same chemotherapy, analyzed all samples in one batch across four assay modes, and made inference at the patient level. Sampling, including draws after breakthrough reactions, fit into routine desensitization care.

The main limitation is size: with 10 allergic patients and four controls, precision and power were limited across all analyses. All participants were women, all allergic patients had gynecologic cancer, and three of four controls had squamous cell carcinoma, so the groups differed in cancer type and the findings may not apply to men or to other cancers. Eleven of the 14 patients were White. Differences in premedication, infusion procedures, and specimen quality constrain allergy-specific interpretation, and visual hemolysis assessment limits interpretation of metabolites released from blood cells. Draws after breakthrough reactions were not timed relative to symptom onset or rescue treatment, which limits interpretation of transient mediators such as histamine; untargeted LC-MS/MS also gave only relative histamine peak areas, not concentrations from a validated quantitative assay. Most features lacked Level 1 identification, and paired changes involving imputed zeros depend on the substituted value.

Untargeted metabolomics of plasma collected during routine carboplatin and paclitaxel desensitization detected premedication exposure and broad pre-to-post infusion changes, mostly shared with tolerant controls, but no allergy-specific signature. Larger prospective studies with timed reaction sampling, premedication-matched comparators, and standardized specimen handling are needed to determine whether such profiling can identify markers of breakthrough reactions or of response to add-on therapies such as omalizumab.

## Supporting information

Online Repository

## Data Availability

The data are not publicly available because they contain clinical information on a small number of patients. Requests for de-identified data can be directed to the senior author, Cosby A. Stone Jr (Vanderbilt University Medical Center), and are subject to institutional approval.

## Acknowledgments

We thank the Vanderbilt Center for Innovative Technology metabolomics core, the clinical research staff of the chemotherapy desensitization program, and the participants in this study.

## Abbreviations

CI: confidence interval
FDR: false discovery rate
HILIC: hydrophilic interaction liquid chromatography
LC-MS/MS: liquid chromatography-tandem mass spectrometry
LPE: lysophosphatidylethanolamine
LysoPC: lysophosphatidylcholine
Q-TOF: quadrupole time-of-flight
RPLC: reversed-phase liquid chromatography
SM: sphingomyelin

