## Supplementary material for "Plasma Metabolomics During Chemotherapy Desensitization: A Pilot Study": Online Repository

### Methods

#### Clinical records and samples

The study comprised 76 draws from 14 patients, collected between October 2024 and October 2025 (Figure E1). Encounters are identified by patient number and sampled-encounter order; for example, A05-3 is patient A05's third sampled encounter. Patient codes (A01 to A10, allergic; T01 to T04, tolerant controls) were assigned for this report and are not linked to medical record or study enrollment numbers. The 39 encounters comprised 37 complete pairs and two post-only encounters (A05-3 and A09-1).

#### Desensitization protocol and premedication

A published report from the same desensitization program describes pharmacy-prepared three-bag, 13-step desensitization with relative dilutions of 1:100, 1:10, and 1:1. Premedication comprised cetirizine 10 mg or fexofenadine 180 mg twice daily, famotidine 20 mg twice daily, and montelukast 10 mg daily, from two days before through two days after desensitization, with standard chemotherapy premedication when indicated.[E1] Some infusions after a breakthrough reaction used longer steps. Sampled desensitization encounters followed this protocol and premedication.

#### Specimen handling

Plasma was obtained during automated cell separation (RoboSep; 72 draws) or density-gradient separation in Ficoll barrier tubes (four draws); one pair from patient A10 used different separation methods at the pre- and post-infusion draws. Fourteen post-infusion draws (12 allergic, two control) were processed on the next calendar day, including the on-treatment post-infusion draws from A07-7 and A09-5; all pre-infusion draws were processed on the day of collection.

Hemolysis was noted in 26 draws (Figure E2), including seven of the nine breakthrough post-infusion draws and all six draws from patient A08. Among omalizumab recipients, patient A09's off-treatment pair (A09-2) was hemolyzed and Ficoll processed, whereas all six of the patient's on-treatment draws were RoboSep processed without noted hemolysis; patient A07's on-treatment pair A07-7 was hemolyzed at both draws.

#### Assay and feature processing

Mass spectrometry-based metabolomic and lipidomic analyses were performed in the Vanderbilt Center for Innovative Technology using previously developed methods,[E2] with deproteinization followed by solid-phase extraction for metabolites and liquid-liquid extraction for lipids. Individual plasma samples (60 $\mu$L) and quality control (QC) pooled reference samples, with added isotope-labeled internal standards, were prepared, dried in vacuo, reconstituted, and analyzed by mass spectrometry. The QC pooled reference samples were made by mixing equal volumes of the individual plasma samples before sample preparation. They were used for column conditioning and retention time alignment, to assess instrument reproducibility throughout the sample sets, and to determine individual sample acceptance from the recovery of the isotope-labeled internal standards.

Individual samples were prepared using previously described methods for metabolites[E3] and lipids.[E4,E5,E6] Metabolomic analyses used liquid chromatography-tandem mass spectrometry (LC-MS/MS; Thermo Scientific Vanquish UPLC coupled to a Q Exactive HF Orbitrap) with previously described methods: hydrophilic interaction liquid chromatography (HILIC) in positive and negative ion modes[E3,E7,E8,E9] and reversed-phase liquid chromatography (RPLC) in negative ion mode.[E10,E11] Lipidomic analyses used an LC-MS/MS system

(Agilent 1290 Infinity LC coupled to an Agilent 6560 Q-TOF) in negative ion mode with previously described methods.[E4]

Metabolomic and lipidomic raw data were imported, processed, normalized to all compounds, and reviewed in Progenesis QI 3.0 (Waters Corporation). All MS and MS/MS sample runs were aligned against a QC pooled reference run for each method (HILIC positive, HILIC negative, RPLC negative, and lipid negative), and unique ions (retention time and $m/z$ pairs) were deconvoluted into features (called compounds in Progenesis). Metabolite annotations were assigned from retention time, accurate mass (error below 5 ppm), isotope distribution similarity, and MS/MS fragmentation spectrum matching against reference databases, including an in-house library and the Human Metabolome Database.[E2,E12] Lipid annotations were assigned from accurate mass (error below 10 ppm), isotope distribution similarity, and, when available, MS/MS spectrum matching, using MS-DIAL database searching, Lipid Annotator,[E13] and LipidMatch.[E14] Annotation confidence followed a five-level scheme adapted from the Metabolomics Standards Initiative: Level 1, confirmed or validated identification; Level 2, putative identification; Level 3, tentative characterization; Level 4, molecular formula; and Level 5, unique or unknown feature.[E2,E15] Across all modes, instrument reproducibility (coefficient of variation [CV] of 5% or less for all metabolite and lipid methods) and sample preparation reproducibility (CV of 18% or less for lipids and 21% or less for metabolites) met the core's quality control criteria.

#### Feature filtering and imputation

Statistical analyses used the normalized peak areas, with zero values (not detected) treated as missing. A feature was retained if no more than 20% of its values across the 76 study samples were missing and its CV (standard deviation divided by mean) across the seven injections of the pooled quality control sample was at most 30%. The 5,212 remaining zeros were replaced by half the feature's smallest positive peak area. Filtering and imputation used all study samples without reference to clinical labels, and no further normalization was applied.

The four modes (in the order used throughout: HILIC positive, HILIC negative, RPLC negative, and lipid negative) contributed 3,551, 3,230, 2,115, and 1,339 input features, of which 2,491, 3,017, 1,858, and 1,280 were retained. Feature identifiers (for example, F1451) number features within each mode. Forty-six input features had Level 1 annotations, of which 45 were retained. Because a feature can represent an adduct, fragment, isotope, or separate chromatographic detection of the same molecule, features sharing a compound annotation were analyzed individually rather than merged.

#### Statistical definitions

Let $Y_{ijkt} > 0$ be the normalized, imputed peak area for patient $i$, encounter $j$, feature $k$, and time point $t$, with $t = 0$ pre and $t = 1$ post. The paired change and its patient mean are

$$d_{ijk} = \log_2 Y_{ijk1} - \log_2 Y_{ijk0}, \qquad \bar{d}_{ik} = \frac{1}{m_i} \sum_{j \in E_i} d_{ijk},$$

where $E_i$ contains patient $i$'s eligible encounters and $m_i = |E_i|$. The primary estimate is

$$\widehat{\theta}_k = \frac{1}{10} \sum_{i \in A} \bar{d}_{ik} - \frac{1}{4} \sum_{i \in C} \bar{d}_{ik},$$

where $A$ and $C$ denote allergic and tolerant-control patients. Positive values indicate a more positive paired change in allergic patients.

##### Patient-label permutation and intervals

The feature-level two-sided statistic was $|\widehat{\theta}_k|$. The exact $P$ value was the proportion of $\binom{14}{4} = 1{,}001$ label assignments, including the observed assignment, with an absolute contrast at least as large as observed. The minimum attainable $P$ value is 1/1,001.[E16] For $m$ features per mode, Benjamini-Hochberg adjustment[E17] rejects the $r$ hypotheses with smallest $P$ values when $P_{(r)} \leq 0.05\, r/m$. Having $r \geq m/(1{,}001 \times 0.05)$ features with $P = 1/1{,}001$ is sufficient, but not necessary, for rejection. Thus, within the respective modes, having at least 50, 61, 38, or 26 features tied at $P = 1/1{,}001$ would suffice for all tied features in that mode to meet the 5% false discovery rate threshold; the observed counts at $P = 1/1{,}001$ were 7, 17, 22, and 2. The test is exact when patient summaries are exchangeable under the null hypothesis, an assumption that unequal encounter counts across patients can weaken.[E18]

Welch inference used the group sample variances $s_A^2, s_C^2$ of patient means and sizes $n_A, n_C$:

$$\mathrm{SE}_k = \sqrt{s_A^2/n_A + s_C^2/n_C}, \qquad \nu_k = \frac{(s_A^2/n_A + s_C^2/n_C)^2}{(s_A^2/n_A)^2/(n_A - 1) + (s_C^2/n_C)^2/(n_C - 1)}.$$

Pointwise 95% confidence intervals (CIs) were $\widehat{\theta}_k \pm t_{.975,\nu_k}\mathrm{SE}_k$.[E19] These intervals apply to individual features, without multiplicity adjustment.

##### Repeated-measures sensitivity

For each retained feature, we fitted a random-intercept model to the primary-comparison encounters:

$$d_{ijk} = \beta_{0k} + \beta_{1k} I(i \in A) + b_{ik} + \epsilon_{ijk}, \quad b_{ik} \sim N(0, \tau_k^2), \quad \epsilon_{ijk} \sim N(0, \sigma_k^2).$$

Here, $\beta_{1k}$ is the allergic-minus-control difference and $b_{ik}$ is the patient random intercept. We used restricted maximum likelihood in `nlme` 3.1-169 with approximate $t$ inference for the fixed effect.[E20] Patient variance was near zero (intraclass correlation below $10^{-6}$) for 508, 932, 372, and 132 features; model and patient-mean estimates correlated at 0.989 to 0.995 (Figure E4).

##### Exploratory empirical-Bayes and Storey sensitivities

Two-group linear models were fitted per feature to the same 14 patient means using `limma` 3.68.5, applying robust empirical-Bayes variance moderation both with and without an abundance-dependent variance trend. The trend covariate was the mean of pre- and post-infusion $\log_2$ abundances, with equal patient weights. An unmoderated pooled-variance $t$ test was included to separate the effect of variance moderation from that of assuming equal group variances. Storey $q$ values were computed from Welch $P$ values in `qvalue` 2.44.0 with fixed $\lambda$ of 0.5 and the package smoother. We also refitted both empirical-Bayes variants restricting to features without imputed values across all 76 draws (1,990, 2,609, 1,646, and 1,159 features). Multiplicity adjustment was performed within each mode and method (Table E4, showing selections at 5% and 10% false discovery rates [FDRs]).

In the primary feature set, the single empirical-Bayes candidate at a 10% FDR was HILIC-negative feature F1451, a Level 4 (molecular formula) annotation, with a between-group difference of +2.36 $\log_2$. Its patient-mean change variances were 0.24 in allergic patients and 1.05 in controls, and its Benjamini-Hochberg-adjusted Welch $P$ value was .82. In HILIC-negative refits excluding features with imputed values, robust moderation selected five features at a 10% FDR (F1451, three other Level 4 annotations, and one Level 3 annotation), and the abundance-trend variant selected three (F1451 and two other Level 4 annotations). None met the 5% FDR threshold, and the selections depended on the method.

#### Global statistics

Between-group global tests used squared Euclidean distances between group means after centering features and dividing by their standard deviations across patient summaries. Calibration used the same 1,001 label assignments. Within-group pre/post tests enumerated $2^{10} = 1{,}024$ allergic and $2^4 = 16$ control sign patterns, multiplying each patient's mean paired-change vector by one shared sign, for $n$ patients in the group, with feature scales $r_k = \sqrt{n^{-1}\sum_i \bar{d}_{ik}^2}$ and statistic $T = \sum_{k:r_k>0}(n^{-1}\sum_i \bar{d}_{ik}/r_k)^2$. Full sign reversal leaves the statistic unchanged, giving minimum two-sided $P$ values of 2/1,024 for allergic patients and 2/16 for controls. Holm adjustment covered 20 global tests: five questions (allergic versus control change, pre-infusion difference, carboplatin-restricted change, allergic pre/post, and control pre/post) in four modes.[E21] Within-allergic feature tests applied these patient sign flips one feature at a time, with Benjamini-Hochberg adjustment within mode.

##### Analysis populations

In every paired encounter, the pre- and post-infusion draws had the same omalizumab exposure status (on or off treatment). Table E1 lists the encounters in each comparison. Two supplementary comparisons used the same controls: breakthrough allergic pairs versus controls (S1) and non-breakthrough allergic pairs versus controls (S2). Within each mode, all S1 and S2 feature tests formed one family for Benjamini-Hochberg adjustment. The pre-infusion comparison used pre draws from the primary-comparison encounters. For the within-person breakthrough contrast, patient A04 contributed A04-1 versus A04-2, A04-3, and A04-4, and patient A07 contributed A07-3 versus A07-1, A07-2, and A07-4. The omalizumab comparison included six on-treatment pairs and five off-treatment pairs from two patients.

#### Results

##### Pre-infusion differences and premedication

Table E2 shows montelukast and cetirizine detection (Level 2 annotations) by group and draw. No individual feature differed between groups before infusion at a 5% within-mode false discovery rate (Figure E3).

##### Pre-to-post change within allergic patients

Within-allergic feature sign-flip permutation tests yielded 129, 609, 78, and 254 features at a 5% within-mode FDR (1,070 total). Of these, 88.4%, 76.5%, 57.7%, and 94.5% changed in the same direction as the mean control change. Thirty-six had confirmed (Level 1) or putative (Level 2) annotations (Table E3), including montelukast (decrease), tetrahydrobiopterin, queuine, oxoglutaric acid, and several nucleosides and organic acids.

#### Allergic versus tolerant patients

The breakthrough-versus-control comparison (S1; seven breakthrough pairs, five patients) and the non-breakthrough-versus-control comparison (S2; 20 non-breakthrough allergic pairs, seven patients) used the same four control pairs from four patients. Neither yielded any feature at a 5% false discovery rate (FDR) after joint within-mode adjustment. Comparisons of carboplatin-allergic patients with controls (six allergic patients; 20 versus four pairs) and of carboplatin-allergic with paclitaxel-allergic patients (six versus three patients; 20 versus four pairs, excluding the patient allergic to both) also yielded no feature at a 5% FDR. Mode-wide global $P$ values for all 20 tests are in Table E6.

Figure E4 compares the patient-mean and random-intercept analyses, Figure E6 shows the bile-acid annotations, and Figure E7 shows the principal components of patient-mean paired changes. Table E5 gives the numbers of pairs and patients retained under each specimen-quality restriction. None of these restrictions yielded a feature at a 5% within-mode false discovery rate, nor did stricter missing-value filters (0%, 5%, or 10% per feature).

The 5,212 substituted zeros occurred in 1,242 retained features; the remaining 7,404 required no substitution. Figure 2 in the main text plots zero values in a separate row rather than at substituted intensities, and Figure E5 shows all retained features and marks those with substituted zeros. All 42 features outside the horizontal range displayed in Figure 3, B, in the main text had at least one zero value among the primary-comparison encounters. Twenty-three had nominal exact $P < .05$, and three had nominal Welch $P < .05$; none met the 5% within-mode FDR threshold. When the analysis was restricted to encounters with positive signal at both draws, 38 of these 42 features had data from at least two patients per group, and 35 of the 38 retained the sign of the original between-group difference.

#### Omalizumab recipients

Figure E8 shows on-minus-off paired changes in the two omalizumab recipients. Zero counts varied across draws: in HILIC-positive mode, the median was 28 zeros per draw, compared with 151 and 144 in the on-treatment post draws from encounters A09-4 and A09-5. These two encounters account for two of the three on-treatment values behind patient A09's larger median absolute paired change (Figure 4, C, in the article).

#### Complete feature estimates

Feature-level results for all retained features are available from the corresponding author on request: group mean paired changes, between-group differences with pointwise Welch 95% CIs, exact and Welch $P$ values, and adjusted $P$ values, with the corresponding mixed-model, empirical-Bayes, supplementary, pre-infusion, culprit-drug, breakthrough, omalizumab, bile-acid, and within-allergic results.

E15. Sumner LW, Amberg A, Barrett D, Beale MH, Beger R, Daykin CA, et al. Proposed minimum reporting standards for chemical analysis: Chemical Analysis Working

Group (CAWG) Metabolomics Standards Initiative (MSI). Metabolomics 2007; 3:211–21

E16. Phipson B, Smyth GK. Permutation P-values should never be zero: calculating exact P-values when permutations are randomly drawn. Stat Appl Genet Mol Biol 2010; 9:Article 39

E17. Benjamini Y, Hochberg Y. Controlling the false discovery rate: a practical and powerful approach to multiple testing. J R Stat Soc Series B Methodol 1995; 57:289–300

E18. Winkler AM, Ridgway GR, Webster MA, Smith SM, Nichols TE. Permutation inference for the general linear model. Neuroimage 2014; 92:381–97

E19. Welch BL. The generalization of ‘Student’s’ problem when several different population variances are involved. Biometrika 1947; 34:28–35

E20. Laird NM, Ware JH. Random-effects models for longitudinal data. Biometrics 1982; 38:963–74

E21. Holm S. A simple sequentially rejective multiple test procedure. Scand J Stat 1979; 6:65–70

**Table E1: Encounters included in the main comparisons**

| Population | Encounter identifiers |
|---|---|
| Breakthrough allergic pairs (S1) | A01-1; A04-1; A05-1, A05-2, A05-4; A07-3; A09-2 |
| Non-breakthrough allergic pairs (S2) | A02-1 to A02-5; A03-1 to A03-3; A04-2 to A04-4; A06-1; A07-1, A07-2, A07-4; A08-1 to A08-3; A10-1, A10-2 |
| On omalizumab | A07-5 to A07-7; A09-3 to A09-5 |
| Off omalizumab, same recipients | A07-1 to A07-4; A09-2 |
| Controls | T01-1; T02-1; T03-1; T04-1 |
| Post-only, excluded from paired analyses | A05-3; A09-1 |

The primary comparison comprises the breakthrough and non-breakthrough allergic pairs (27 pairs, 10 patients) and the control pairs (four pairs, four patients), all off omalizumab. The within-person breakthrough and omalizumab comparisons include overlapping patients and are descriptive. Encounter identifiers give patient and sampled-encounter order. S1, supplementary comparison 1 (breakthrough versus control); S2, supplementary comparison 2 (non-breakthrough versus control).

**Table E2: Detection of two Level 2 premedication annotations by group and draw**

| Compound | Group | Draw | Draws | Detected | Fraction |
|---|---|---|---|---|---|
| Montelukast | Allergic | Pre | 33 | 33 | 1.00 |
| Montelukast | Allergic | Post | 35 | 34 | 0.97 |
| Montelukast | Control | Pre | 4 | 0 | 0.00 |
| Montelukast | Control | Post | 4 | 0 | 0.00 |
| Cetirizine | Allergic | Pre | 33 | 31 | 0.94 |
| Cetirizine | Allergic | Post | 35 | 31 | 0.89 |
| Cetirizine | Control | Pre | 4 | 2 | 0.50 |
| Cetirizine | Control | Post | 4 | 2 | 0.50 |

Detection means a peak area above zero before imputation. Both annotations used hydrophilic interaction liquid chromatography with positive ionization.

#### Table E3: Level 1 and Level 2 annotations of within-allergic feature changes

| Annotation | Mode | Feature | Confidence level | Allergic change | Control change | Adjusted *P* |
|---|---|---|---|---|---|---|
| EtherPC(34:3) | Lipid negative | F0066 | 2 | −0.09 | −0.10 | .020 |
| Glucuronate | HILIC negative | F0022 | 2 | 1.16 | 1.88 | .028 |
| 4-Methylumbelliferone sulfate | HILIC negative | F0127 | 2 | −0.97 | −1.54 | .028 |
| Nopalinic acid | HILIC negative | F0110 | 2 | 0.85 | −0.02 | .028 |
| Glycerylphosphoryl-ethanolamine | HILIC negative | F0020 | 2 | 0.73 | 0.03 | .028 |
| Temocaprilat | HILIC negative | F0126 | 2 | −0.64 | −0.47 | .028 |
| Oxoglutaric acid | HILIC negative | F0035 | 2 | 0.61 | 0.24 | .028 |
| Sorbose | HILIC negative | F0068 | 2 | 0.45 | −0.01 | .028 |
| Gluconic acid | HILIC negative | F0050 | 2 | 0.43 | 0.56 | .028 |
| Uric acid | HILIC negative | F0005 | 1 | −0.20 | 0.10 | .028 |
| Dehydroascorbate | HILIC negative | F0067 | 2 | 0.64 | −0.77 | .031 |
| Orotidine | HILIC negative | F0059 | 2 | −0.43 | −0.98 | .031 |
| N-acetylalanine | HILIC negative | F0058 | 2 | 0.36 | 0.34 | .031 |
| N-acetylleucine | HILIC negative | F0101 | 2 | 0.81 | −0.07 | .037 |
| 2’-deoxyinosine | HILIC negative | F0017 | 2 | 0.66 | 0.51 | .037 |
| Taurine | HILIC negative | F0004 | 1 | 0.35 | −0.26 | .037 |
| Montelukast | HILIC positive | F0138 | 2 | −1.84 | 0.00 | .038 |
| Tetrahydrobiopterin | HILIC positive | F0033 | 2 | 1.33 | 0.62 | .038 |
| Queuine | HILIC positive | F0090 | 2 | 1.27 | 0.45 | .038 |
| D-alpha-aminobutyric acid | HILIC positive | F0066 | 2 | 0.83 | −0.34 | .038 |
| N-acetylglutamate | HILIC negative | F0010 | 1 | 0.49 | 0.56 | .042 |
| Rhamnose | HILIC negative | F0061 | 2 | 0.41 | 1.78 | .042 |
| Creatinine | HILIC negative | F0048 | 2 | −0.23 | 0.53 | .042 |
| EtherPE(38:4) | Lipid negative | F0099 | 2 | −0.36 | −0.28 | .044 |
| EtherPE(36:6) | Lipid negative | F0030 | 2 | −0.26 | −0.14 | .044 |
| PC(40:5) | Lipid negative | F0086 | 2 | 0.23 | 0.06 | .044 |
| PC(38:5) | Lipid negative | F0047 | 2 | 0.21 | 0.10 | .044 |
| EtherPE(34:3) | Lipid negative | F0063 | 2 | −0.19 | −0.16 | .044 |
| Oxoglutaric acid | RPLC negative | F0007 | 2 | 0.78 | −0.04 | .047 |
| Uridine | RPLC negative | F0008 | 2 | −0.53 | −0.51 | .047 |
| Ipratropium bromide | HILIC negative | F0104 | 2 | 3.57 | 6.37 | .048 |
| Coumestrol | HILIC negative | F0075 | 2 | −1.22 | −0.45 | .048 |
| Monoethyl malonic acid | HILIC negative | F0049 | 2 | 0.54 | −0.18 | .048 |
| Alpha-aminoadipate | HILIC negative | F0046 | 2 | 0.30 | −0.11 | .048 |
| L-glutamic acid | HILIC negative | F0025 | 2 | 0.29 | 0.03 | .048 |
| LPE(20:4) | Lipid negative | F0139 | 2 | 0.61 | −0.18 | .049 |

All 36 features met a 5% within-mode false discovery rate in the within-allergic sign-flip test and are sorted by adjusted *P*. Changes average patient-mean paired $\log_2$(post/pre) changes from primary-comparison encounters. Feature identifiers are numbered within each mode, so the same identifier can appear in two modes. Putative (Level 2) annotations are library matches supported by both precursor mass and MS/MS fragmentation data; the montelukast control change is 0 because the feature was not detected in control draws. HILIC, hydrophilic interaction liquid chromatography; RPLC, reversed-phase liquid chromatography; LPE, lysophosphatidylethanolamine; PC, phosphatidylcholine; PE, phosphatidylethanolamine.

**Table E4: Features selected by method and threshold in the primary comparison**

| Method | HILIC positive | HILIC negative | RPLC negative | Lipid negative |
|---|---|---|---|---|
| Exact patient-label permutation, BH | 0 / 0 (.24) | 0 / 0 (.15) | 0 / 22 (.08) | 0 / 0 (.51) |
| Welch patient-mean $t$, BH | 0 / 2 (.09) | 0 / 0 (.27) | 0 / 2 (.07) | 0 / 0 (.23) |
| Random-intercept mixed model, BH | 0 / 0 (.41) | 0 / 0 (.25) | 0 / 0 (.48) | 0 / 0 (1.00) |
| Pooled-variance $t$, BH | 0 / 0 (.23) | 0 / 0 (.15) | 0 / 0 (.18) | 0 / 0 (.45) |
| Robust empirical Bayes, BH | 0 / 0 (.24) | 0 / 1 (.09) | 0 / 0 (.18) | 0 / 0 (.24) |
| Robust empirical Bayes with abundance trend, BH | 0 / 0 (.21) | 0 / 1 (.07) | 0 / 0 (.15) | 0 / 0 (.48) |
| Welch $t$, Storey $q$ ($\lambda = 0.5$) | 0 / 2 (.08) | 0 / 0 (.22) | 0 / 2 (.07) | 0 / 0 (.23) |
| Welch $t$, Storey $q$ (smoother) | 0 / 2 (.08) | 0 / 0 (.22) | 0 / 2 (.07) | 0 / 0 (.23) |

Entries give feature counts at 5% / 10% adjusted thresholds; parentheses give the minimum adjusted value. Analyses use the same 14 patients: patient means except for the mixed model, which uses all 31 encounters. Adjustment is within mode. The two Welch selections at 10% in HILIC positive are montelukast (F0138, Level 2) and an unannotated feature (F1840); the two in RPLC negative include traumatic acid (putative, Level 2) and an unannotated feature (F0957). Both Storey variants select the same four features. The empirical-Bayes selection is a Level 4 (molecular formula) annotation (F1451). BH, Benjamini-Hochberg; HILIC, hydrophilic interaction liquid chromatography; RPLC, reversed-phase liquid chromatography.

**Table E5: Specimen-quality restrictions in the primary comparison**

| Restriction | Allergic pairs | Patients | Control pairs | Patients |
|---|---|---|---|---|
| All eligible pairs | 27 | 10 | 4 | 4 |
| Same hemolysis status at both draws | 21 | 10 | 4 | 4 |
| No hemolysis at either draw | 14 | 8 | 3 | 3 |
| RoboSep processing at both draws | 25 | 9 | 4 | 4 |
| Both draws processed on collection day | 17 | 9 | 2 | 2 |

Eligible encounters are re-averaged within patient per restriction. Same-day processing uses calendar dates.

**Table E6: Mode-wide global tests**

| Question | HILIC positive | HILIC negative | RPLC negative | Lipid negative |
|---|---|---|---|---|
| Allergic versus control paired change (primary) | .147 (1.00) | .054 (.755) | .093 (1.00) | .569 (1.00) |
| Pre-infusion difference | .001 (.020) | .018 (.270) | .170 (1.00) | .143 (1.00) |
| Carboplatin-allergic versus control paired change | .157 (1.00) | .071 (.929) | .167 (1.00) | .448 (1.00) |
| Pre-to-post change, allergic patients | .002 (.037) | .002 (.037) | .002 (.037) | .004 (.063) |
| Pre-to-post change, controls | .125 (1.00) | .375 (1.00) | .125 (1.00) | .125 (1.00) |

Entries give exact $P$ values with Holm-adjusted values across all 20 tests in parentheses. Between-group tests enumerated 1,001 patient-label assignments (210 for the carboplatin restriction); within-group tests enumerated 1,024 allergic and 16 control sign patterns, with minimum two-sided $P$ values of 2/1,024 and 2/16. HILIC, hydrophilic interaction liquid chromatography; RPLC, reversed-phase liquid chromatography.

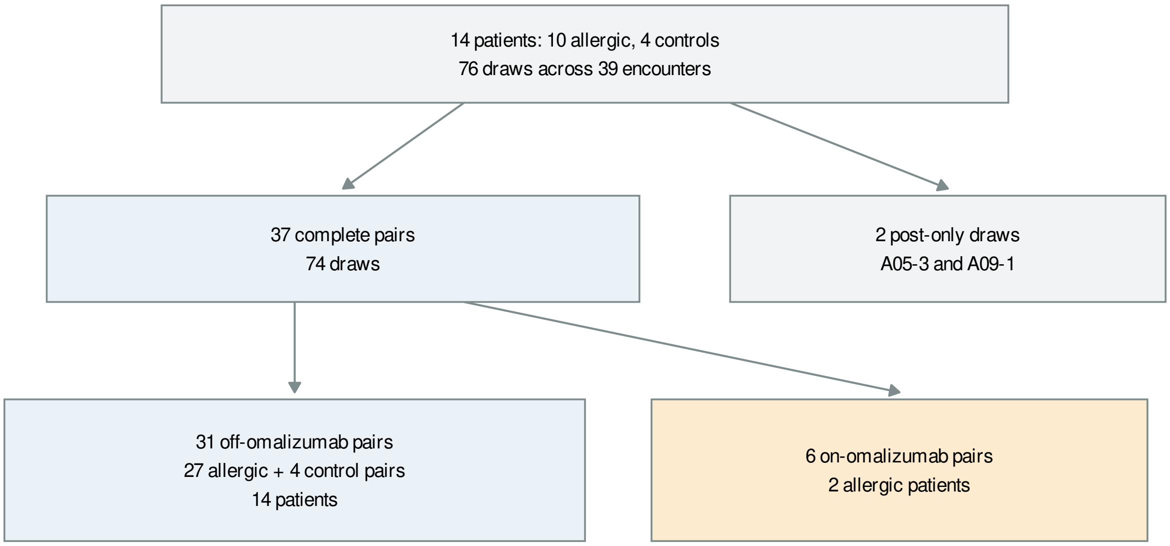


**Figure E1: Cohort and specimen flow.** Draws, encounters, and patients from enrollment to analyzed pairs, including post-only encounters A05-3 and A09-1. Omalizumab branches count on-treatment and off-treatment encounters from the same two recipients.

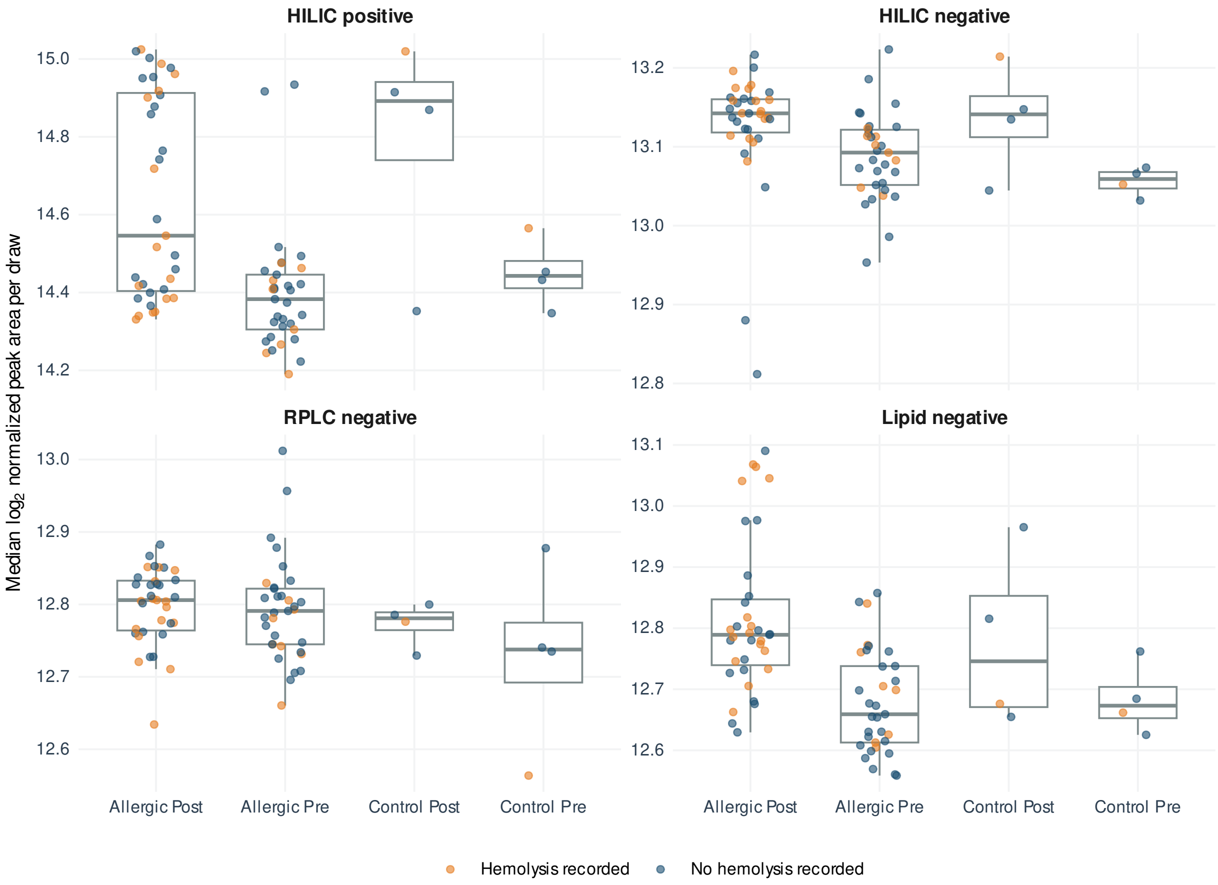


**Figure E2: Median signal per draw by group, time point, and recorded hemolysis.** Each point shows one draw's median $\log_2$ normalized peak area across retained features in one mode. Boxes summarize draws.

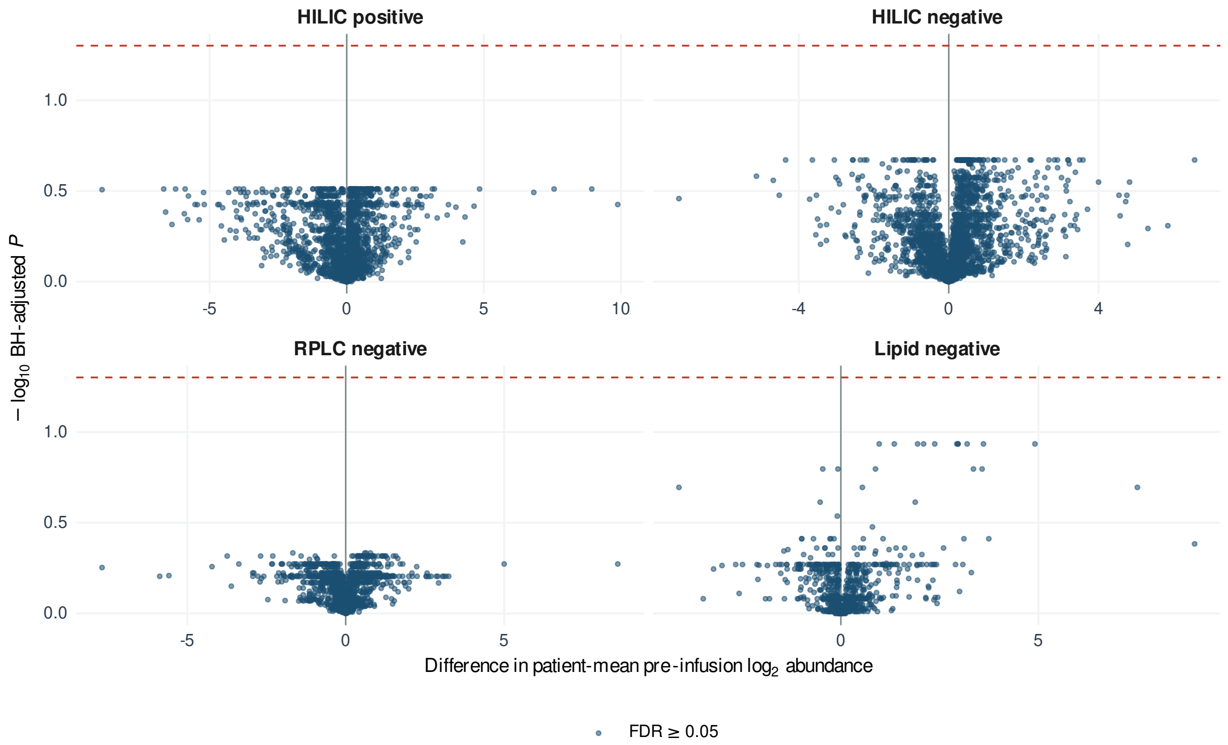


**Figure E3: Pre-infusion allergic-minus-control differences for all retained features.** Differences in patient-mean pre-infusion $\log_2$ abundance plotted against within-mode Benjamini-Hochberg-adjusted exact $P$ values. The dashed line marks .05.

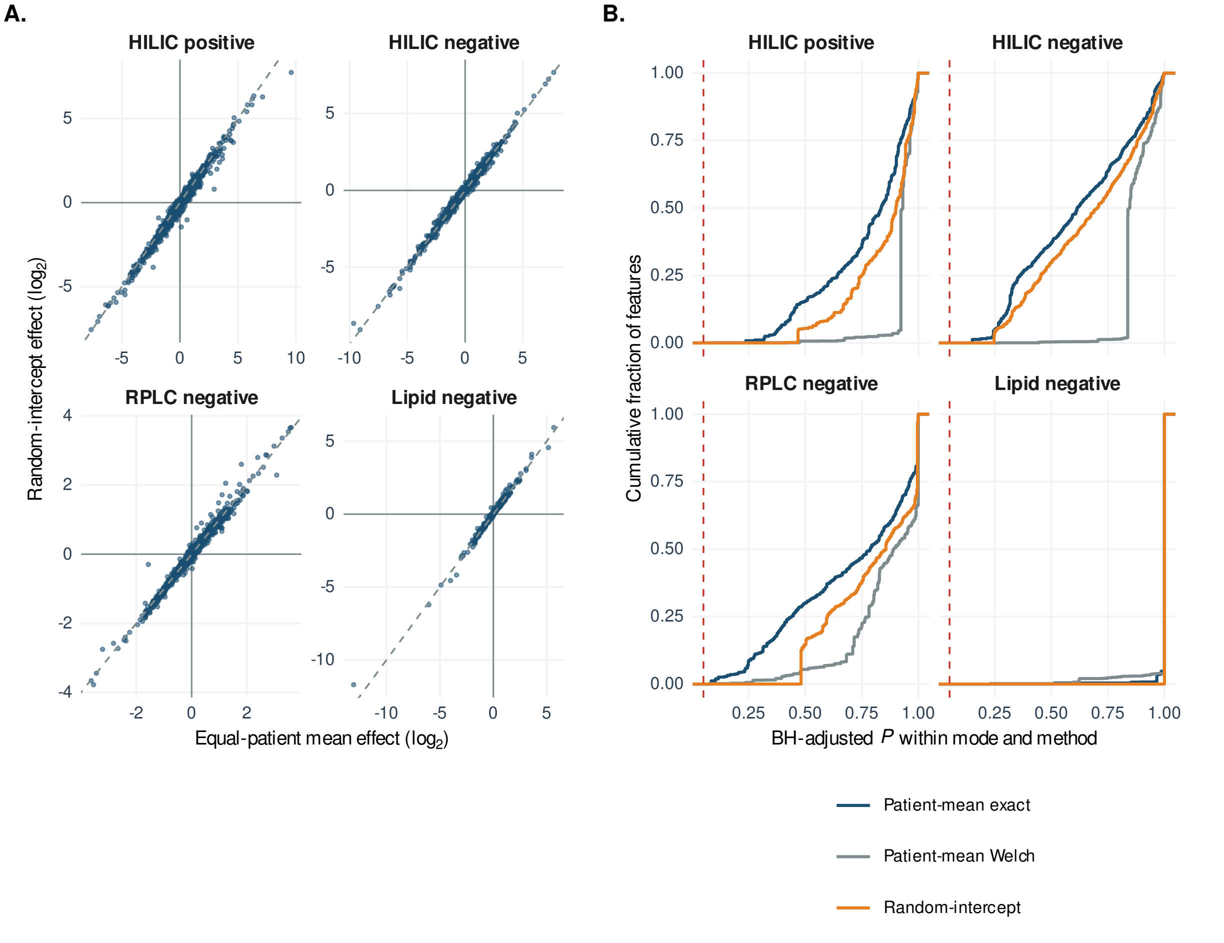


**Figure E4: Patient-mean and mixed-model estimates and adjusted *P* values.** A. Patient-mean and random-intercept effect estimates for retained features; dashed diagonals mark equal estimates. B. Cumulative distributions of Benjamini-Hochberg-adjusted *P* values for exact patient-label, Welch patient-mean, and random-intercept analyses; the red line marks .05.

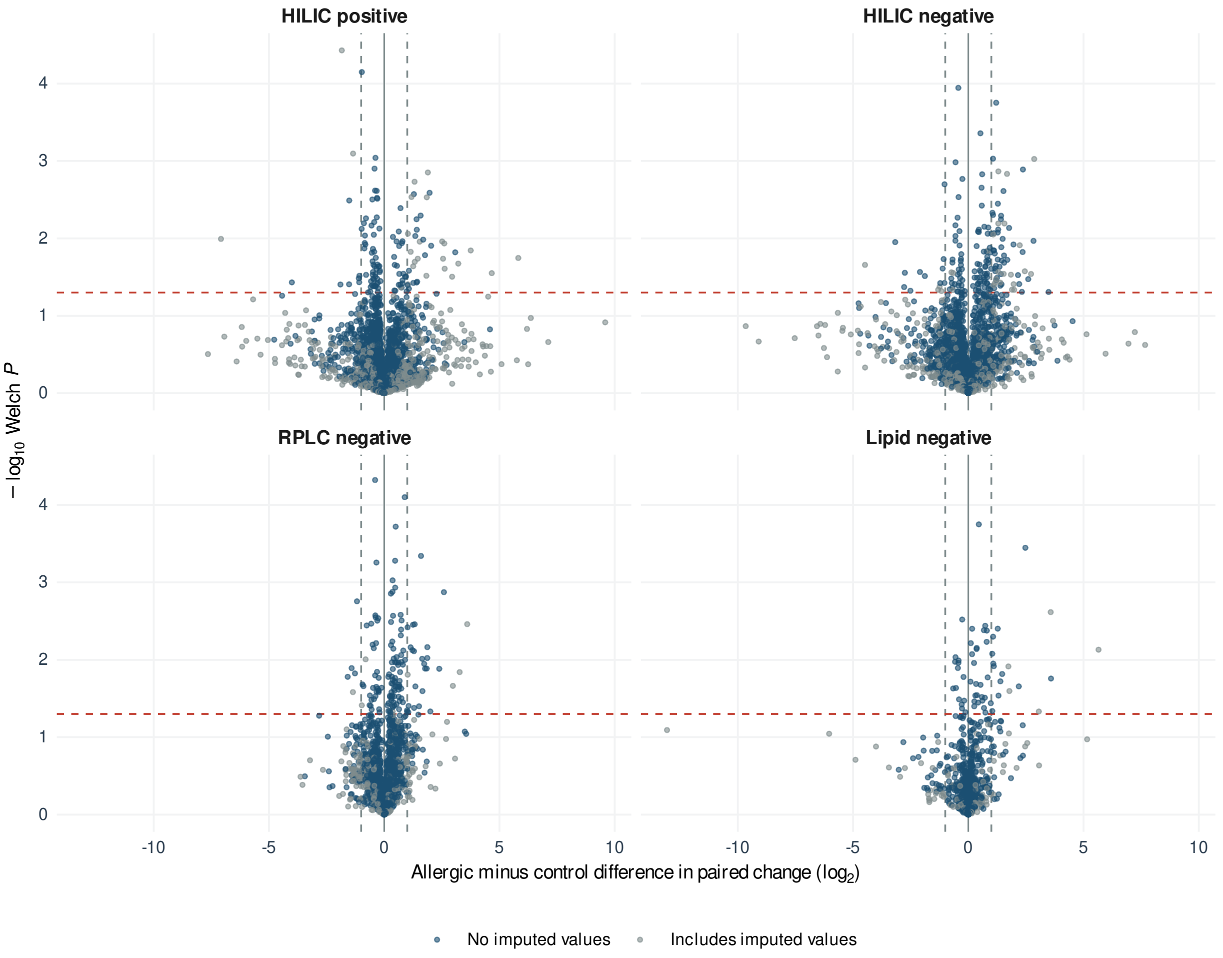


**Figure E5: Full-range primary comparison and substituted zeros.** All 8,646 retained features, including the 42 outside the range of Figure 3, B. Gray points indicate at least one substituted zero among the 76 draws. Dashed vertical lines mark twofold differences in paired change; the dashed horizontal line marks nominal Welch $P = .05$.

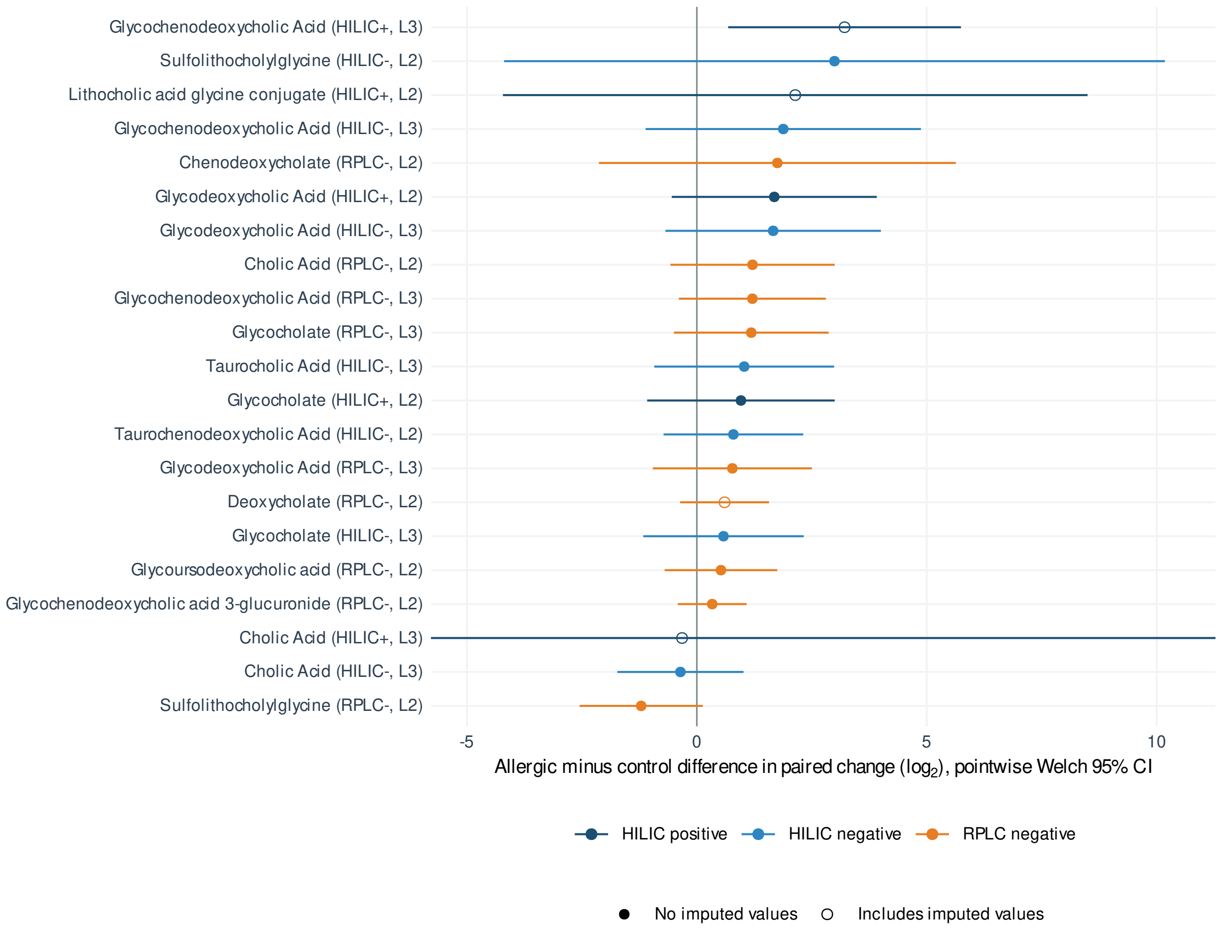


**Figure E6: Bile-acid features in the primary comparison.** All 21 retained features whose top-ranked annotation was a bile acid or bile-acid conjugate (each having at most two candidate annotations). Points show allergic-minus-control differences in patient-mean paired change with pointwise Welch 95% CIs; open symbols indicate features with at least one imputed zero value. The interval for cholic acid (HILIC positive, Level 3) extends beyond the display range.

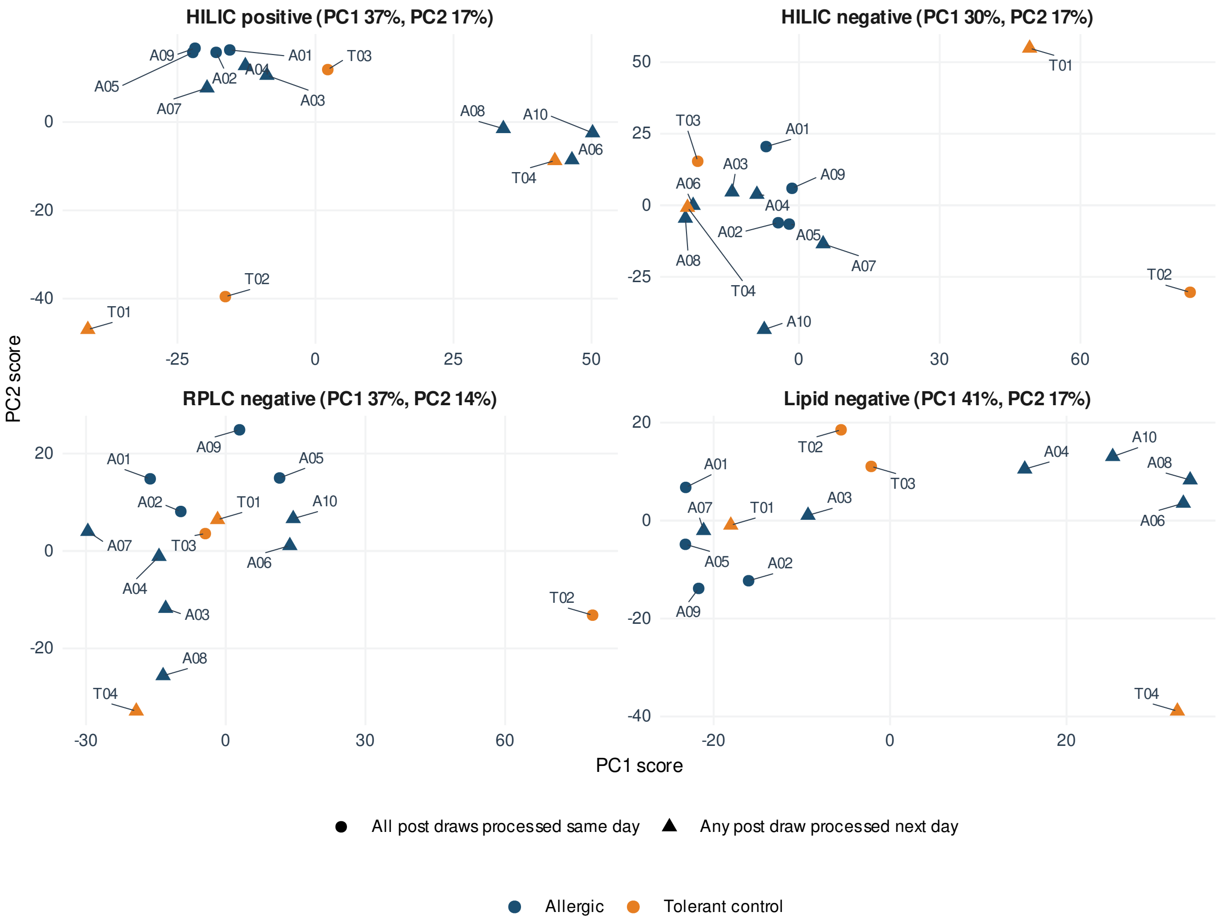


**Figure E7: Principal components of patient-mean paired changes.** Each point represents one patient in the primary comparison. Percentages give the variance captured by each component. Shape indicates whether any of the patient's post draws were processed on the next calendar day.

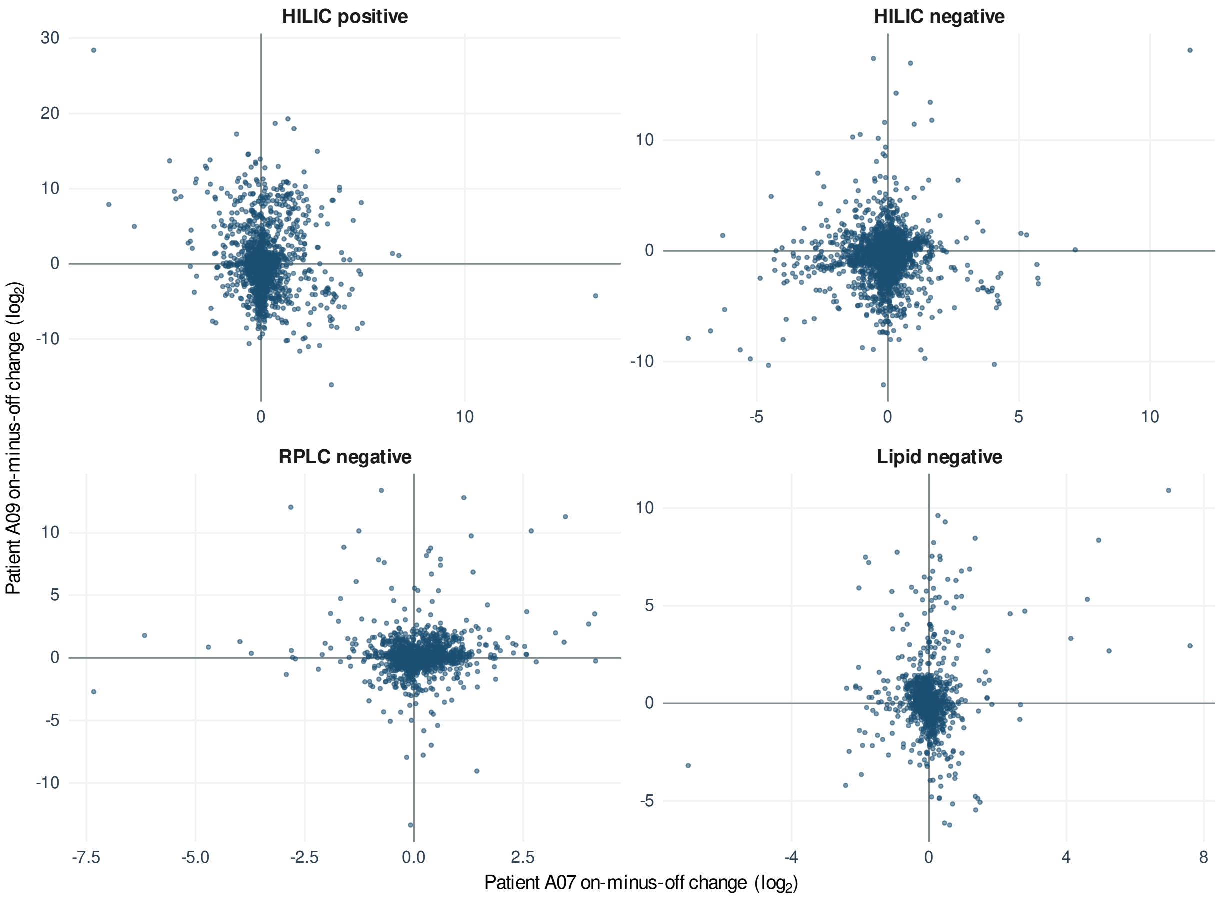


**Figure E8: On-minus-off paired changes in the two omalizumab recipients.** Each point represents one retained feature. Axes show each patient's mean paired $\log_2$ change on treatment minus off treatment, including changes involving substituted zero values.
